# Intracameral Antibiotic Prophylaxis and Surgical Expertise: Key Determinants in Endophthalmitis After Cataract Surgery

**DOI:** 10.1101/2025.05.19.25324616

**Authors:** Vinicius Campos Bergamo, Luis Filipe Nakayama, Nilva Simeren Bueno de Moraes, Ivan Maynart Tavares, Ana Luisa Hofling-Lima, Maurício Maia

**Affiliations:** Retina Division, Department of Ophthalmology, Escola Paulista de Medicina, Hospital São Paulo, Universidade Federal de São Paulo, São Paulo, Brazil; Glaucoma Division, Department of Ophthalmology, Escola Paulista de Medicina, Hospital São Paulo, Universidade Federal de São Paulo, São Paulo, Brazil; Cornea and External Diseases Division, Department of Ophthalmology, Escola Paulista de Medicina, Hospital São Paulo, Universidade Federal de São Paulo, São Paulo, Brazil; Laboratory of Ocular Microbiology Division, Department of Ophthalmology, Escola Paulista de Medicina, Universidade Federal de São Paulo, São Paulo, Brazil

## Abstract

**Background/Objectives:** Postoperative endophthalmitis is a rare but severe complication of cataract surgery. Intracameral moxifloxacin prophylaxis has been widely adopted, but concerns remain regarding bacterial resistance and clinical outcomes. This study aimed to evaluate the impact of intracameral moxifloxacin prophylaxis on endophthalmitis incidence, microbiological patterns, antibiotic resistance, and visual outcomes in a university hospital setting, with comparison to a private hospital cohort.

**Subjects/Methods:** This retrospective cohort study analyzed 21,178 cataract surgeries performed at a university hospital (2014–2023). The incidence of endophthalmitis, microbiological profiles, resistance patterns, and visual acuity outcomes (LogMAR) were assessed. Intracameral moxifloxacin prophylaxis was introduced in 2019. A comparative analysis was conducted with 19,360 surgeries from a private hospital. Fisher’s exact test, Mann-Whitney U test, and Joinpoint regression were used for statistical analysis.

**Results:** Endophthalmitis incidence at the university hospital was 0.109% (23/21,178 surgeries), significantly higher than the 0.021% incidence in the private hospital (p=0.002). Post-prophylaxis, infection rates declined from 0.219% in 2016 to 0.042% in 2023 (p=0.003). *Staphylococcus epidermidis* predominated (52.6%), and moxifloxacin resistance remained stable (pre: 45.5%; post: no increase).

**Conclusions:** Intracameral moxifloxacin significantly reduced endophthalmitis incidence without increasing bacterial resistance. Post-prophylaxis infection rates aligned with national surveillance benchmarks (e.g., SES-SP threshold of 0.07%), reinforcing the efficacy of this preventive strategy in a public university setting. The worsening of final visual acuity post-prophylaxis underscores the need for continued clinical vigilance. Ongoing microbiological surveillance remains essential.

## Introduction

Postoperative endophthalmitis is a rare but severe intraocular infection that remains one of the most feared complications following cataract surgery, with potential for profound visual impairment and irreversible blindness. (1) Despite its low incidence, ranging from 0.04% to 0.3% (2–7), its devastating consequences demand strict preventive strategies and prompt intervention.

Historically, improvements in surgical techniques, surgical equipment and prophylactic protocols have contributed to a gradual decline in infection rates, yet the condition continues to pose a clinical challenge, particularly in light of emerging antimicrobial resistance and shifts in microbiological profiles (8,9)

Multiple patient-related and surgical factors influence the risk of postoperative endophthalmitis. Advanced age, diabetes mellitus, prolonged operative time, posterior capsule rupture, and poor intraoperative wound integrity have been identified as key risk factors. (10–12)

Intracameral antibiotic prophylaxis, particularly with moxifloxacin, has gained acceptance globally due to its demonstrated efficacy in reducing infection rates. (13–15) Following international trends, Federal University São Paulo - Hospital São Paulo - Ophthalmology Department in Brazil introduced routine intracameral moxifloxacin (Vigamox®, Alcon Laboratories) prophylaxis in January 2019.

The aim of this study was to retrospectively evaluate the impact of intracameral moxifloxacin prophylaxis on postoperative endophthalmitis incidence, microbiological patterns, bacterial resistance profiles, and visual outcomes in cataract surgeries performed from January 2014 to December 2023. Additionally, a comparative analysis was conducted between the university public hospital and a private facility to assess the influence of institutional factors on infection rates and clinical outcomes.

## Materials and Methods

This retrospective cohort analysis included data from all cataract surgeries performed between January 2014 and December 2023 at the Ophthalmology Department of the Federal University of São Paulo (UNIFESP), a Brazilian tertiary university hospital (Hospital São Paulo). Additionally, data from cataract surgeries performed in 2023 at the Ophthalmologic Hospital of Brasília (HOB-OPTY) were also collected. The study was approved by the university’s Ethics Committee (protocol number 0060/2018), and the requirement for informed consent was waived due to its retrospective design.

Clinical, microbiological, antibiotic susceptibility, and visual acuity data (measured in LogMAR) were extracted and analyzed. Data were obtained from the Ocular Microbiology Laboratory of the Department of Ophthalmology, Federal University of São Paulo, as well as from electronic medical records. Routine prophylactic intracameral administration of preservative-free moxifloxacin (Vigamox®, Alcon Laboratories) at a dose of 0.1 mL was implemented in January 2019.

Importantly, all other aseptic and antiseptic protocols remained consistent throughout the study period. These included preoperative skin antisepsis with 10% povidone-iodine (PVP-I), instillation of 5% PVP-I in the conjunctival sac, eyelash isolation using sterile drapes, individualized operating rooms, and adherence to national surgical safety standards.

Microbiological identification and susceptibility testing were conducted following standard laboratory protocols. Statistical analyses included descriptive statistics to summarize categorical and continuous variables. Fisher’s exact test was used to assess associations between categorical variables, while the Mann–Whitney U test was applied to compare non-normally distributed continuous variables. For normally distributed variables, means were compared using Student’s *t*-test. Temporal trends were evaluated using joinpoint regression analysis. A significance level of α = 0.05 was adopted for all statistical tests.

## Results

Between January 2014 and December 2023, a total of 21,178 cataract surgeries were performed at the university hospital, with 23 confirmed cases of postoperative endophthalmitis, corresponding to an overall incidence of 0.109% (1.09 cases per 1,000 surgeries). Before the introduction of intracameral moxifloxacin prophylaxis in 2019, annual incidence rates fluctuated, reaching peaks of 0.219% in 2016 and 0.205% in 2017. Following prophylaxis implementation, a progressive decline was observed, with infection rates dropping to 0.066% in 2022 and 0.042% in 2023. Statistical analysis using Joinpoint regression confirmed this downward trend as statistically significant (Annual Percentage Change: −15.81%; 95% CI: −24.94 to −5.58; p=0.003), reinforcing the effectiveness of intracameral moxifloxacin in reducing postoperative infections. (Figures 1 and 2) Table 1 presents the annual distribution of endophthalmitis cases, including the total number of surgeries performed and the corresponding incidence rates for each year.

**Figure 1.**
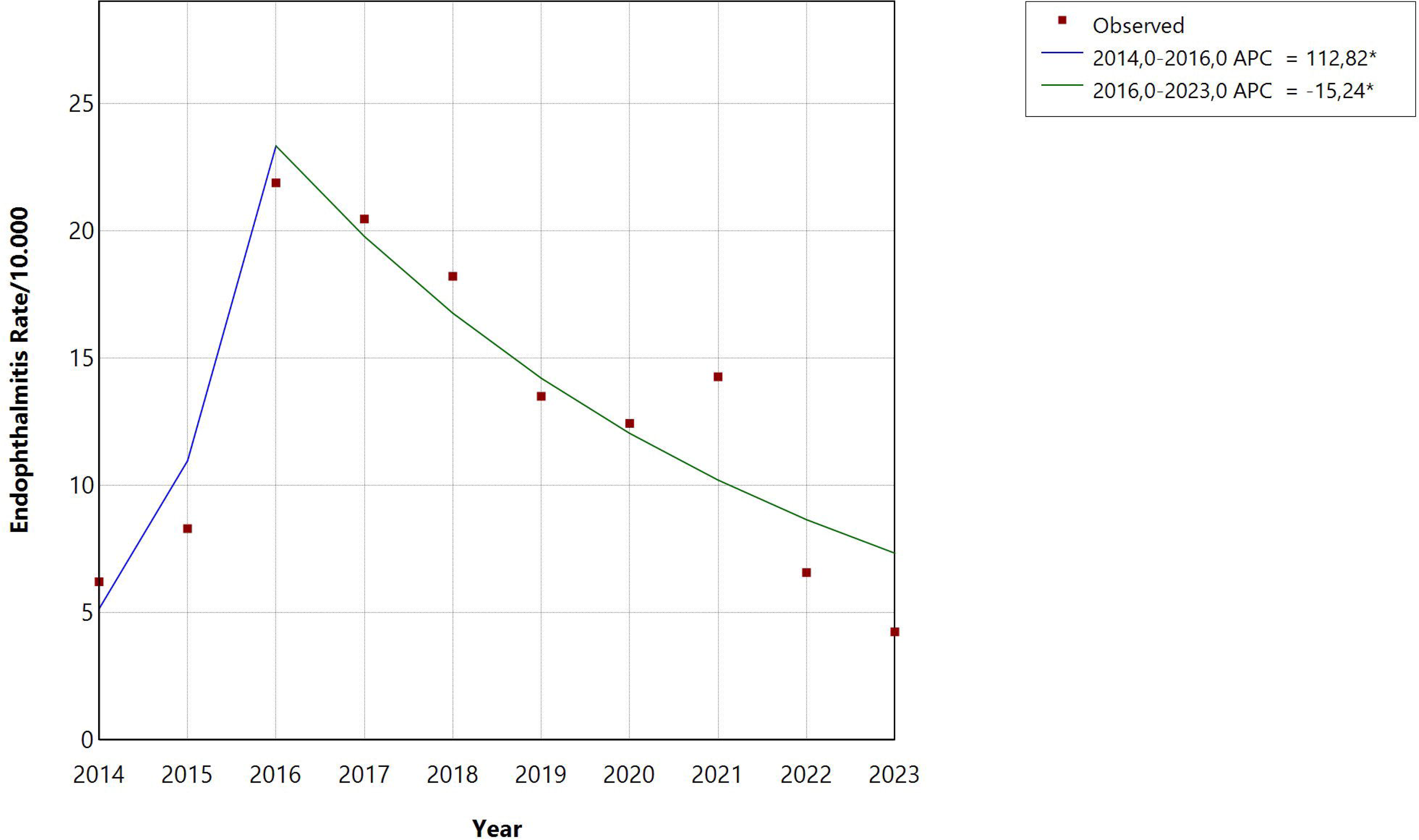
Annual incidence of postoperative endophthalmitis after cataract surgery (2014–2023). A declining trend was observed following the introduction of intracameral moxifloxacin in 2019.

**Figure 2.**
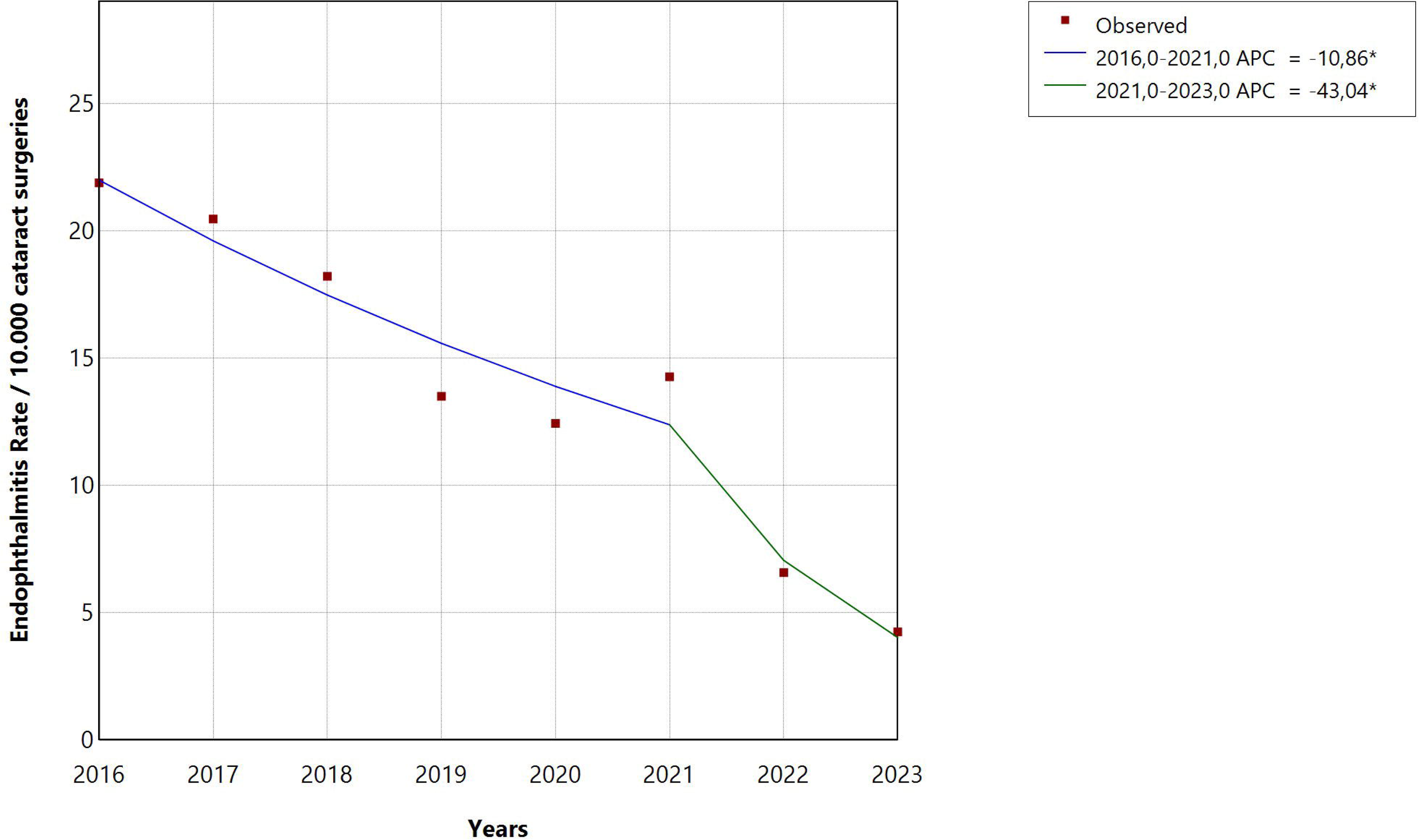
Joinpoint regression analysis of endophthalmitis incidence (2014–2023). A significant annual reduction of –15.81% (95% CI: –24.94 to –5.58; p = 0.003) was observed after prophylaxis implementation.

Statistical analyses, including Fisher’s exact test and independent Student’s t-test, demonstrated no significant differences in baseline patient characteristics between the pre- and post-prophylaxis groups, including age, diabetes status, and history of prior surgeries. These findings are summarized in Table 2.

Throughout the study period, Gram-positive bacteria predominated, particularly *Staphylococcus epidermidis* and coagulase-negative staphylococci, which together accounted for 52.6% of all positive cultures. An increase in bacterial diversity was observed post-prophylaxis, with isolated cases of Gram-negative infections, including *Pseudomonas aeruginosa*. These findings are summarized in Table 3.

Regarding antibiotic resistance patterns, prior to prophylaxis implementation (2014– 2018), moxifloxacin resistance was detected in 45.5% of isolates, while 66.6% of strains were resistant to oxacillin, and moderate to high resistance to cephalosporins was observed.

Vancomycin remained fully effective (100% sensitivity) throughout the study period. In the post-prophylaxis period (2019–2023), no statistically significant increase in moxifloxacin resistance was detected. Vancomycin retained its efficacy, and while a slight increase in Gram-negative isolates was noted, there was no clear evidence of a shift in overall pathogen virulence. These findings are summarized in Table 4.

Final visual acuity (VA) outcomes varied among patients, with LogMAR values distributed across a broad range. Following the introduction of intracameral moxifloxacin prophylaxis, there was no statistically significant difference in VA between pre- and post-prophylaxis groups (Mann-Whitney U test, p = 0.670). Severe visual impairment (LogMAR ≥2.8) was observed in some cases, despite prophylactic measures. These findings indicate that while prophylaxis reduced the overall incidence of endophthalmitis, a subset of patients still experienced poor visual outcomes.

A comparative analysis was conducted using data from a private hospital, where 19,360 cataract surgeries were performed within the year of 2023. Only four cases of postoperative endophthalmitis were recorded in this setting, resulting in a significantly lower incidence rate of 0.021%.

## Discussion

This study provides strong evidence that routine intracameral moxifloxacin prophylaxis significantly reduces the incidence of postoperative endophthalmitis following cataract surgery.

In 2023, our institution achieved a postoperative endophthalmitis rate of 0.042%, which falls below the 0.07% threshold adopted by regional surveillance programs such as the São Paulo State Health Secretariat (SES-SP). (16) Although the Brazilian Health Regulatory Agency (ANVISA) does not define a strict national benchmark, it mandates monthly reporting and monitoring of endophthalmitis cases as part of its national infection surveillance system. Our findings meet these surveillance goals, underscoring both the effectiveness of intracameral prophylaxis and institutional adherence to national standards.

The implementation of intracameral moxifloxacin marked a significant shift in infection control practices, addressing concerns over infection rates observed in previous years. While variability in rates was evident before its adoption, the introduction of this prophylactic measure coincided with a notable downward trend, reinforcing the efficacy of intracameral antibiotic prophylaxis in mitigating postoperative infections.

The observed reduction in endophthalmitis incidence aligns with previous studies, particularly the landmark trial by the European Society of Cataract and Refractive Surgeons (ESCRS), which demonstrated a fourfold decrease in infection rates with intracameral cefuroxime. (17,18) Similar findings have been reported with moxifloxacin, especially in high-volume surgical centers. (13,14,19) Our results further validate its effectiveness in a university-based public hospital setting, where the baseline infection risk may be inherently higher due to factors such as increased surgical volume, resident participation, and variations in aseptic technique. (20,21)

Consistent with previous literature, microbiological analysis confirmed that Gram-positive bacteria, particularly *Staphylococcus epidermidis* and coagulase-negative staphylococci, were the predominant pathogens in both pre- and post-prophylaxis periods. (22–24) The introduction of intracameral moxifloxacin did not significantly alter the microbiological profile, although a slight increase in Gram-negative isolates, including *Pseudomonas aeruginosa*, was noted post-prophylaxis.

Culture positivity rates declined following the introduction of intracameral prophylaxis. Before 2019, the average culture positivity rate was 81.25%, indicating that most infections were microbiologically confirmed. However, after the introduction of prophylaxis, positivity rates declined to 62.5%, suggesting a reduced bacterial load at the time of infection or an increase in culture-negative cases. This reduction may indicate that intracameral prophylaxis effectively lowered bacterial inoculum, leading to milder infections or more challenging microbiological detection. Concerns have been raised regarding the potential for intracameral antibiotic prophylaxis to induce bacterial resistance, particularly with widespread fluoroquinolone use. (25,26)

However, our findings indicate that this risk has not yet materialized. Pre-prophylaxis, moxifloxacin resistance was detected in 45.5% of isolates, with significant resistance also noted for oxacillin (66.6%) and cephalosporins. In the post-prophylaxis period, no statistically significant increase in moxifloxacin resistance was observed, and vancomycin retained 100% efficacy. These findings suggest that intracameral moxifloxacin remains an effective prophylactic agent, with no significant increase in bacterial resistance observed during the study period. Despite theoretical concerns regarding the widespread use of intracameral fluoroquinolones, continued microbiological surveillance is essential to detect potential resistance shifts over time.

Although there appeared to be a clinical trend towards improved final visual acuity outcomes following the introduction of intracameral moxifloxacin prophylaxis, statistical analysis (Mann-Whitney U test, p = 0.670) showed no significant difference between pre- and post-prophylaxis groups, consistent with previous literature. (27) This finding suggests that while intracameral moxifloxacin effectively reduced overall endophthalmitis incidence, its impact on the severity of individual infections, as measured by final visual acuity, was less clear.

The persistence of severe cases despite the introduction of prophylaxis highlights critical considerations. One possible explanation is that prophylactic measures may significantly reduce overall bacterial inoculum but might not entirely prevent infections caused by particularly virulent or resistant strains. Additionally, a reduced overall incidence may inadvertently lower clinical suspicion, potentially leading to delayed diagnosis and intervention, as both physicians and patients may become less alert to symptoms of endophthalmitis. These factors underscore the importance of maintaining rigorous postoperative monitoring protocols and continuous patient and healthcare provider education to ensure early detection and intervention.

Another potential factor influencing visual acuity outcomes could be the timing and approach to management. Delayed intervention or initial underestimation of infection severity might have contributed to suboptimal visual results in some cases. Future studies should explore whether early and aggressive interventions, such as prompt pars plana vitrectomy (PPV), could improve visual outcomes, particularly in breakthrough infections occurring despite prophylaxis.

A noticeable finding in this study was the five-fold lower infection rate observed in the private hospital cohort compared to the university hospital (0.021% vs. 0.109%), despite the implementation of similar prophylactic protocols. This disparity suggests that factors beyond prophylaxis, such as surgical experience, institutional protocols, and infection control measures, may contribute significantly to postoperative infection risk. At the university hospital, cataract surgeries were performed by both attending ophthalmologists (fellows) and residents in training, whereas in the private hospital, all procedures were conducted by fully trained surgeons. Prior research has suggested that surgeries performed by residents may have a slightly higher complication rate, including endophthalmitis, likely due to prolonged surgical times, increased intraocular manipulation, and greater variability in technique. (20) Additionally, high-volume university hospitals, particularly those publicly funded, often face resource constraints and higher patient turnover, potentially affecting surgical duration, surgical efficiency, and postoperative surveillance. In contrast, private hospitals may benefit from more controlled surgical environments, stricter operating room protocols, more standardized surgical practices, and enhanced postoperative monitoring, all of which could contribute to lower infection rates.

The relevance of intracameral antibiotic prophylaxis in reducing postoperative endophthalmitis has also been demonstrated in other public healthcare settings. A recent Brazilian study by Salha et al. (28) evaluated the introduction of intracameral cefuroxime in a tertiary public hospital where cataract surgeries were predominantly performed by residents. Following implementation, the incidence of endophthalmitis dropped from 0.27% to 0%, with no reported adverse effects. These findings emphasize not only the effectiveness and safety of intracameral prophylaxis, but also its critical role in public teaching hospitals, where inherent challenges such as variability in surgical expertise and limited resources may increase infection risk.

Beyond institutional differences, socioeconomic disparities may also play a role in postoperative infection risk. Patients from lower socioeconomic backgrounds may face barriers to accessing high-quality healthcare services, including delayed surgical care, limited access to postoperative follow-up, and difficulty in adhering to medical recommendations. Studies have suggested that poorer living conditions, reduced health literacy, and financial constraints may contribute to suboptimal perioperative hygiene and lower compliance with postoperative care protocols, potentially increasing susceptibility to infectious complications, including endophthalmitis. (21) Additionally, patients treated in public hospitals often experience longer wait times for surgery and a higher burden of systemic comorbidities, such as diabetes, which has been associated with an increased risk of postoperative infections. (1,10,11,29)

Although this study did not directly assess the impact of socioeconomic and institutional factors, the significant difference in infection rates between the public university hospital and the private hospital highlights the need for further investigation into how socioeconomic determinants, surgical training programs, and institutional infection control policies influence postoperative infection risk and visual outcomes. These findings reinforce the efficacy of intracameral moxifloxacin in reducing infection rates but emphasize the necessity of maintaining a high index of suspicion for endophthalmitis.

Early recognition, prompt diagnosis, and aggressive management remain crucial, as delays continue to significantly impact visual outcomes. (30) Further research into refining prophylactic and therapeutic strategies, particularly for identifying cases at higher risk of severe outcomes, remains essential.

This study has important clinical implications. Routine intracameral moxifloxacin should remain the standard of care in cataract surgery, given its demonstrated ability to significantly reduce endophthalmitis rates without evident resistance selection. However, the five-fold lower infection rate observed in private hospitals suggests that optimizing infection control strategies in academic settings may further enhance patient safety. Training programs should emphasize surgical asepsis, early infection recognition, and refined intraoperative techniques to minimize infection risks among resident-performed surgeries. Moreover, despite a declining incidence of endophthalmitis, clinicians should remain highly vigilant, as delayed recognition may contribute to worse visual outcomes.

This study has limitations inherent to its retrospective design. The reduced number of microbiological samples in the post-prophylaxis period limits the ability to detect subtle resistance trends. Furthermore, the differences between hospital settings (private vs. university) may involve confounding variables not accounted for in our analysis. Future prospective, multicenter studies should explore the impact of surgeon experience, institutional infection control measures, and host factors on postoperative endophthalmitis incidence and clinical outcomes.

In conclusion, routine intracameral moxifloxacin prophylaxis significantly reduced postoperative endophthalmitis incidence without increasing bacterial resistance. However, the observed discrepancy in infection rates between university and private hospitals highlights the critical role of surgical experience and institutional infection control policies. Additionally, it underscores the need for ongoing clinical vigilance, emphasizing early recognition and aggressive management of residual cases. Continued microbiological surveillance is essential to detect potential future resistance trends and to further refine prophylactic strategies in cataract surgery.

## Data Availability

All data produced in the present study are available upon reasonable request to the authors

## Author Contribution Statement

VCB, LFN, NSBM, IMT, MSDQC, ALHL, and MM all made substantial contributions to the conception and design of the study, acquisition and interpretation of data, and critical revision of the manuscript for important intellectual content. All authors approved the final version of the manuscript and agree to be accountable for all aspects of the work in ensuring the accuracy and integrity of any part of the study.

## Summary Box

### What was known before

- Postoperative endophthalmitis is a rare but severe complication of cataract surgery, with variable incidence depending on surgical setting and preventive measures.
- Intracameral antibiotic prophylaxis, especially with moxifloxacin or cefuroxime, has been shown to reduce the incidence of endophthalmitis in large-volume surgical centers.
- Differences in infection rates between institutions may reflect disparities in surgical training, experience, and infection control protocols.

### What this study adds

- Intracameral moxifloxacin prophylaxis significantly reduced the incidence of endophthalmitis in a public university hospital setting over a 10-year period.
- No increase in moxifloxacin resistance was observed after routine use, supporting its safety and microbiological stability.
- A five-fold lower infection rate in a private hospital cohort suggests that surgical expertise and institutional factors play a major role in infection prevention, beyond pharmacological measures.

