## Supplementary material for "Intracameral Antibiotic Prophylaxis and Surgical Expertise: Key Determinants in Endophthalmitis After Cataract Surgery": Table 1

Table 1. Annual Incidence of Postoperative Endophthalmitis Following Cataract Surgery (2014–2023)

**Table 1 – Trends in endophthalmitis rates (per 10,000 surgeries), according to the Joinpoint regression model**

| **Tendência 1** | | |  | **Tendência 2** | | |  | **Total** | | |
| --- | --- | --- | --- | --- | --- | --- | --- | --- | --- | --- |
| **Period** | **APC** | **CI95%** |  | **Period** | **APC** | **CI95%** |  | **Period** | **AAPC** | **CI95%** |
| 2014 - 2016 | 108.63 | -10.89 a 388.46 |  | 2016 - 2023 | -15.81* | -24.94 a -5.58 |  | 2014 - 2023 | 3.00 | -12.18 a 20.79 |
| 2016 - 2021 | -11.29* | -17.75 a -4.33 |  | 2021 a 2023 | -41.93 | -68.36 a 6.60 |  | 2016 - 2023 | -21.41* | -29.73 a 12.10 |

APC: Annual Percentage Change; AAPC: Average Annual Percentage Change; CI: Confidence Interval; * Statistically Significant
