## Supplementary material for "Intracameral Antibiotic Prophylaxis and Surgical Expertise: Key Determinants in Endophthalmitis After Cataract Surgery": Table 2

Table 2. Annual Incidence of Postoperative Endophthalmitis Following Cataract Surgery (2014–2023)

| **Year** | **Total Surgeries** | **Endophthalmitis Cases** | **Positive Cultures** | **Endophthalmitis Rate (%)** |
| --- | --- | --- | --- | --- |
| **2014** | 3221 | 1 | 1 | 0,031% |
| **2015** | 2411 | 1 | 1 | 0,041% |
| **2016** | 2284 | 5 | 4 | 0,219% |
| **2017** | 1954 | 4 | 3 | 0,205% |
| **2018** | 2196 | 3 | 2 | 0,137% |
| **2019** | 2222 | 3 | 1 | 0,135% |
| **2020** | 1609 | 2 | 1 | 0,124% |
| **2021** | 1402 | 2 | 1 | 0,143% |
| **2022** | 1522 | 1 | 1 | 0,066% |
| **2023** | 2357 | 1 | 1 | 0,042% |
|  | **21178** | **23** | **16** | **0,109%** |
