## Supplementary material for "Intracameral Antibiotic Prophylaxis and Surgical Expertise: Key Determinants in Endophthalmitis After Cataract Surgery": Table 3

Table 3. Comparison of Baseline Characteristics Between Pre- and Post-Prophylaxis Groups (2014–2018 vs. 2019–2023)

|  | **Years** | | **p** |
| --- | --- | --- | --- |
|  | **2014 to 2018** | **2019 to 2023** |  |
| Age (years), N Mean ± SD | 14 66.64 ± 13.88 | 9 63.11 ± 19.85 | 0.825^a^ |
| VA, N Mean ± SD | 4 1.59 ± 0.98 | 9 1.15 ± 1.13 | 0.310^a^ |
| Culture, n(%) |  |  | 0.363^b^ |
| No | 3/14 (21.4) | 4/9 (44.4) |  |
| Yes | 11/14 (78.6) | 5/9 (55.6) |  |
| PCR, n(%) |  |  | 0.657^b^ |
| No | 10/14 (71.4) | 5/9 (55.6) |  |
| Yes | 4/14 (28.6) | 4/9 (44.4) |  |
| Previous/Combined Glaucoma Surgery, n(%) |  |  | 1.000^b^ |
| No | 12/14 (85.7) | 8/9 (88.9) |  |
| Yes | 2/14 (14.3) | 1/9 (11.1) |  |
| Diabetes, n(%) |  |  | 1.000^b^ |
| No | 10/14 (71.4) | 6/9 (66.7) |  |
| Yes | 4/14 (28.6) | 3/9 (33.3) |  |

p - descriptive level of the Mann-Whitney test (^a^) and Fisher’s Exact test (^b^).

VA – Visual Acuity; PCR – Posterior Capsule Rupture
