## Supplementary material for "Intracameral Antibiotic Prophylaxis and Surgical Expertise: Key Determinants in Endophthalmitis After Cataract Surgery": Table 4

Table 4. Microbiological Profile of Endophthalmitis Cases Before and After Intracameral Moxifloxacin Prophylaxis (2014–2018 vs. 2019–2023)

| **Pre-prophylaxis (2014-18)** | | |  | **Post-prophylaxis (2019-23)** | | |
| --- | --- | --- | --- | --- | --- | --- |
| S. epidermidis | 4 | 36.36% |  | S. coagulase negative | 2 | 28.57% |
| S. coagulase negative | 2 | 18.18% |  | S. aureus | 2 | 28.57% |
| S. aureus | 2 | 18.18% |  | Streptococcus orali (viridans group) | 1 | 14.29% |
| Strepto pneumoniae | 1 | 9.09% |  | Micrococcus luteus | 1 | 14.29% |
| Morganella morganii | 1 | 9.09% |  | Pseudomonas aeruginosa | 1 | 14.29% |
| Anaerobium Gram negative bacillus | 1 | 9.09% |  |  | **7** | **100%** |
|  | **11** | **100%** |  |  |  |  |
