## Supplementary material for "Intracameral Antibiotic Prophylaxis and Surgical Expertise: Key Determinants in Endophthalmitis After Cataract Surgery": Table 5

| **Organism** | **Amikacin** | **Ceftriaxone** | **Ciprofloxacin** | **Gentamicin** | **Moxifloxacin** | **Ofloxacin** | **Oxacillin** | **Tobramycin** | **Vancomycin** |
| --- | --- | --- | --- | --- | --- | --- | --- | --- | --- |
| S. coagulase negative | S | S | S | S | S | S | R | S | S |
| S. epidermidis | S | S | S | R | S | S | S | S | S |
| S. epidermidis | S | R | R | R | R | R | R | R | S |
| S. epidermidis | S | S | R | R | R | R | S | S | S |
| Strepto pneumoniae | N | N | N | N | S | S | S | N | S |
| Bac GN anaerobic | N | N | N | N | N | N | N | N | N |
| S. aureus | S | R | R | S | I | R | R | S | S |
| Morganella morganii | S | S | I | S | R | R | N | S | N |
| S. coagulase negative | S | N | R | S | R | R | R | S | S |
| S. epidermidis | S | N | R | R | I | R | R | R | S |
| S. aureus | S | N | R | S | R | R | R | S | S |
| S. coagulase negative | S | N | S | S | S | S | S | S | S |
| Streptococcus orali (viridans group) | N | S | N | N | N | S | N | N | S |
| S. aureus | S | N | R | S | S | R | S | S | N |
| S. epidermidis | S | N | R | R | R | R | R | R | S |
| Micrococcus luteus + Pseudomonas aeruginosa | N | N | N | N | N | N | N | N | N |
| S. coagulase negative + S. aureus | S | N | R | R | R | N | S | R | N |

Table 5. Antibiotic Susceptibility Patterns of Isolated Microorganisms in Endophthalmitis Cases (Pre- and Post-Prophylaxis Periods)

**Post-Prophylaxis period is highlighted**
